# Prospective case-control study of dengue infection in some malaria and non-malaria patients consulting at the Bertoua Regional Hospital, East-Cameroon

**DOI:** 10.1101/2021.09.03.21263073

**Authors:** Borris Rosnay Galani Tietcheu, Elodie Ndeme Ayangma, Sylvie Wouatedem, Huguette Claire Meke Nguele

## Abstract

In Cameroon, recent studies have documented several cases of dengue infection in some urban areas, but dengue-malaria coinfection malaria was little investigated, especially in the East region. From the 6th July to the 4th September 2020, a case-control study including 50 cases (malaria-positive subjects) and 90 controls (non-malaria subjects) was carried out at the Bertoua Regional Hospital. Participants were prospectively enrolled and administered with a questionnaire to record information such as age, sex, dwelling place, dengue knowledge, and the environment’s quality. Blood specimens were then collected and screened for dengue infection using the NS1/IgG/IgM rapid diagnostic tests and hematological parameters were measured using a MINDRAY-type hemacytometer. Of the malaria patients, 14% (7/50) were tested positive for dengue fever against 66.66% for the controls (60/90). Most malaria patients had a secondary dengue infection (57.14%, 4/7) while most of the non-malaria patients faced a primary infection (61.66%, 37/60). In both groups, women were more exposed than men to dengue and there was a significant association between the dwelling place and dengue seropositivity. Moreover, young subjects <16 years old were significantly more associated with dengue than 16-30 years old (OR=16.24, P=0.042 for cases vs, OR=21, P=0.0001 for controls). The analysis of hematological parameters showed a significant decrease (P<0.001) in platelets in cases compared to control. These results suggest that dengue fewer targets malaria- than non-malaria patients with different serological characteristics in Bertoua city. However, co-infected patients demonstrated a greater clinical vulnerability than monoinfected patients, urging the need for epidemiological surveillance.

## Introduction

Dengue is the most widespread mosquito-borne viral disease throughout the world in particular in tropical areas [1]. According to WHO, the number of dengue cases reported over the last decades grew from 505,430 cases in 2000 to 5.2 million in 2019 worldwide [2]. Some predictions state that 2.25 (1.27–2.80) billion more people will be exposed to dengue from here to 2080 leading the whole population at risk to over 6.1 billion [3]. In Africa, previous studies have documented numerous dengue cases, some of them causing severe epidemics [4,5]. Moreover, dengue fever is endemic in several countries and frequently associated with the etiology of febrile illnesses interfering thereby with certain infectious diseases like malaria. Indeed, both infections are clinically difficult to differentiate as they display similar symptoms [6].

Dengue is caused by the Dengue virus (DENV), a single-stranded positive RNA virus, member of the Flaviviridae family which presently exists under four serotypes (DENV1-4) having each distinct antigenic and genetic features [7]. These serotypes are able to induce two types of dengue namely classic dengue characterized by symptoms such as retroorbital pain, headache, severe myalgias, and severe hemorrhagic dengue fever which causes more serious complications resulting in bleeding and death [1]. DENV is mainly carried and spread to humans by Aedes mosquitoes [8]. Recent evidence suggests that human-driven changes including deforestation, urbanization ineffective vector control, and climate change would have greatly promoted the expansion of this infection to the African continent. [3].

In Cameroon, several studies have reported the presence of dengue infection in urban and rural areas [9–16]. Different dengue serotypes were also characterized in especially DENV-1, DENV-2, and DENV-3 serotypes in asymptomatic [17] and febrile patients [9,14]. Dengue-malaria coinfection was firstly documented in children attending healths centers in Yaoundé [15]. Later, Nkengfou et al found a coinfection rate of 19.5% in infants consulting at the Centre Mère-Enfant in Yaounde [10]. In another study, a malaria-dengue coinfection rate of 13% was documented in the New-Bell district in Douala [14]. More recently, we reported a seroprevalence of 8.95% in a cohort of patients surveyed in the Ngaoundere Regional Hospital in the Adamawa region [18]. However, in the East region of Cameroon, there are currently no data on DENV circulation, to the best of our knowledge. Nevertheless, previous reports gave evidence of the invasion of this region by *Aedes aegypti* and *Aedes albopictus* [19,20] as in other towns such as Douala and Yaounde where dengue infection has been identified. Moreover, several cases of severe malaria have been documented in this region [21]. According to Antonio-Kondjo et al. the East region is a reservoir for malaria transmission in Cameroon and is considered by the Ministry of Health as one of the most affected areas by malaria [22]. Therefore, concurrent dengue and malaria infections are not to be excluded in people living in this area. The aim of this work was to bring data on dengue presence in the Bertoua city and to perform a compared analysis of this infection between malaria and non-malaria patients.

## Materials and Methods

### Study site

This cross-sectional and descriptive study was conducted from 6th July to the 4th September 2020 at the Bertoua Regional Hospital (BRH) (Fig 1).

**Fig 1:**
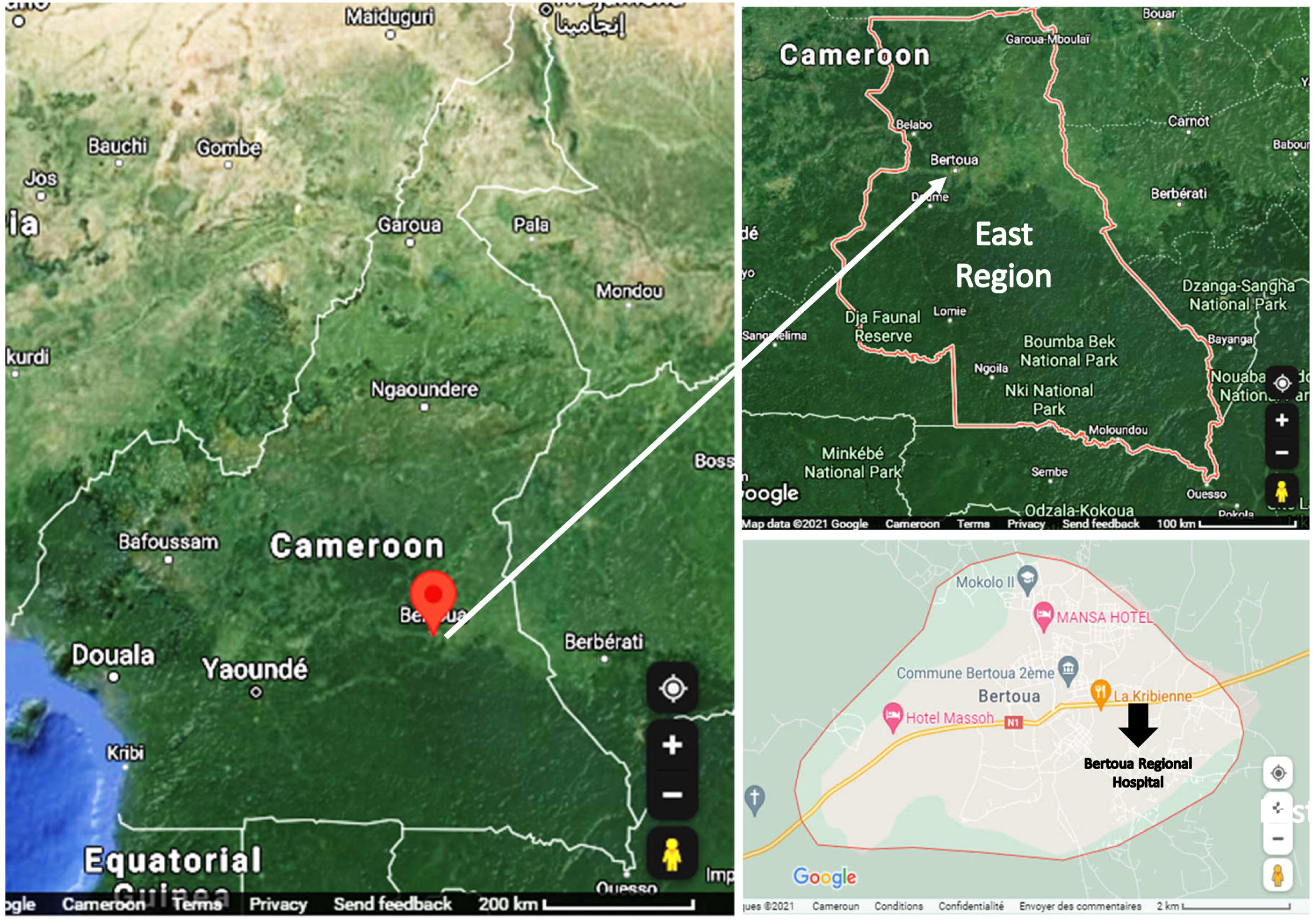
Geographical map showing the location of the Bertoua Regional Hospital.

Bertoua is the capital of the Department of Lom and Djerem and the regional capital of the East, the largest forest region in Cameroon. It is located 350 km from yaoundé with the following geographical coordinates 4 ° 34 ′ 30 ″ north, 13 ° 41 ′ 04 ″ east. It covers 100 km2 with around a whole population estimated to 281139 inhabitants [23]. Its climate is subtropical with three seasons including a large dry season which begins in December and ends in mid-March; a small rainy season which goes from mid-March to mid-May, a big rainy season which goes from mid-September to November. The temperature is high (average between 23-25 ° C) with a peak around 30°C. The average oscillates between 23 and 25 °C [24]. Precipitation is relatively abundant there (1,500 to 2,000 mm of rainfall per year).

### Ethical Considerations

This study protocol was approved by the National Ethics Committee for Research on Human Health (CNRESH) (Ethical Clearance No. 2020/01/58/ CE/CNERSH/SP). Permission for sample collection at BRH was granted by the Regional Delegate for Public Health of the East region (Authorization N°622/L/MINSANTE/SG/DRSPE/BFP). The risks and benefits of the study were explained to each participant, afterwards, a written and signed consent form was obtained from each voluntary subject. Assents and consents for young participants less than 21 years old were given by their parents or legal guardians.

### Study population and inclusion criteria

Have been included in the study, volunteer patients aged between 0-50 years old, and consulting at the BRH. Patients presenting at least two of the following symptoms: headache, asthenia, myalgia, rash, arthralgia, nausea, abdominal pain, vomiting, and loss of appetite and screened positive for malaria infection were recruited as the case group (n=50) while those screened negative for malaria with no evidence of the above-mentionned symptoms were included in the control groups (n=90). All patients were administered a standardized questionnaire to collect socio-demographic data as well as the sex, age, dwelling place, dengue knowledge, and the environment’s quality. Were excluded, patients under malaria treatment, with immunodeficiencies or sexually transmitted diseases.

### Laboratory tests

#### Malaria parasite detection

Malaria screening was carried out using a whole blood thick smear as previously described [25]. Briefly, a drop of freshly collected blood was placed on a degreased slide, then the latter was spread regularly using a vaccinostyle for 2 min. The slide was then inverted and then dried flat on a support for 1 hour at room temperature. The thick drop was then covered abundantly with 3 drops of a 10% Giemsa solution and 2 ml of neutral water. The whole was incubated for 5-10 minutes until completely discolored and then fixed with methyl alcohol. The excess liquid was poured carefully and replaced again with the Giemsa and neutral water mixture for 20 min of incubation. The slide was washed gently with distilled water and then air dried. The thick drop was observed under a microscope at 100x magnification using oil immersion.

#### Dengue diagnosis tests

Dengue screening was carried out using the NS1/IgG/IgM combined diagnostic kit provided by Medsinglong Global Group Co., Limited (Guangzhou, China). It is a one-step rapid test based on the principle of immunochromatography with double-antibody sandwich immunoassay and capture made of two devices: the NS1 antigen test (on the left side) and the IgG/IgM test (at the right side). After adding the sample to the test device, the specimen is absorbed and mixed with the antibody/antigen conjugates. Then, it flows across the pre-coated membrane. When the NS1 antigen levels in the sample are above the detection limit of the test, they combined to the antibody conjugate and then are fixed by the anti-NS1 antibody immobilized in the test region (T) of the device giving a colored band in T which shows a positive result. Regarding the IgG/IgM test, when the dengue-specific antibodies in the specimen are at/above the limit of detection, they bind to the antigen conjugate in the conjugate pad and then are catched by the anti-human IgG or anti-human IgM immobilized in the test region (T1 and T2) of the device giving therefore a positive result in these regions. All tests were carried out following the manufacturer’s instructions. For the NS1 test, three drops of the blood sample (60-80µl) were added in the appropriate well in the cassette device using a dropper, while 10µl of the plasma sample were added in well A followed by 2 drops of the dilution buffer in the B well for the IgG/IgM test. The cassette was let at room temperature for 20 min and the results were interpreted following the manufacturers instructions.

As stated previously [18], the performances of this kit were compared with the Australian Panbio dengue test kit. A sensitivity of 92.22% (166/180) and 97.30% (180/185) were reported with the NS1 antigen and IgG/IgM antibody respectively, while specificity of 100% (170/170) was observed with the NS1antigen and 98.18% (162/165) with the antibodies. Similarly, no cross-reactivity was found with the chikungunya, measles, rubella, typhoid fever, leptospirosis, influenza, hemorrhagic fever with renal syndrome, septicemia, tsutsugamushi disease, hepatitis B and C, HIV, malaria *P. falciparum, P. vivax*, and epidemic cerebrospinal meningitis.

#### Measurement of hematological parameters

The haematological parameters of the study participants were determined using a MINDRAY brand automated hemacytometer. Its principle was based on the counting of the Figd elements of the blood in an automatic way and the measurement of their proportion in the plasma liquid. To perform this test, 5 mL of blood was drawn using a syringe and placed in a tube containing the anticoagulant ethylene diamine tetra acetate (EDTA). The tube was shaken, then placed in the hemacytometer which aspirated 2 mL of this blood. A few minutes after this device displayed the result. The following parameters were evaluated: the number of erythrocytes, leukocytes, platelets, hematocrit, lymphocytes, neutrophils and mean blood volume.

### Statistical analysis

Data collected were processed in excel spreadsheets. Statistical analyzes were conducted using the Graphpad Prism 5.0 software (San Diego, USA). Data were displayed as mean plus or minus standard error to mean, and percentages. Mean comparisons were performed using the two-way analysis of variance followed by a Bonferonni posttest. The χ2 test was used to determinate associations between dengue seropositivity, and the socio-demographic characteristics and the odds ratios (OR) were calculated The significant difference was declared for P <0.05.

## Results

### General characteristics of the study population

The general characteristics of the study population are displayed in Table 1. As shown by this table, women (60%) were more represented than men (40%) in both case and control groups. The mean age of the case group was 25.5 years old while that of the control was 23.83 years old. In cases groups, subjects from 31-50 years were the most represented (40%) while in the control groups, age ranges were balanced enough (33.33%). In the case groups, most participants originated from Ngaikaida (20%) and Monou (14%) neighborhoods while in the controls, they were issued from Tindamba (16.66%) and Ngaikaida (14%). In both groups, a huge majority of the participants reported to live in a risky environment (94% for cases vs 100% for controls) for dengue infection and were not aware of what dengue infection was about (>96%).

**Table 1.** General characteristics of the study population

|  |  | **Cases** | **Controls** |
| --- | --- | --- | --- |
| Gender | Male | 20 (40%) | 36 (40%) |
|  | Female | 30 (60%) | 54 (60%) |
|  | Total | 50 | 90 |
| Age range (years) | [0-15] | 15 (30%) | 30 (33.33%) |
|  | [16-30] | 15 (30%) | 30 (33.33%) |
|  | [31-50] | 20 (40%) | 30 (33.34%) |
|  | Total | 50 | 90 |
| Mean age (years) |  | 25.5 | 23.83 |
| Dwelling place | Enia | 5 (10%) | 7 (7.77%) |
|  | Tigaza | 5 (10%) | 10 (11.11%) |
|  | Ngaikaida | 10 (20%) | 13(14.44%) |
|  | Yademe | 5 (10%) | 9 (10%) |
|  | Mokolo | 2 (4%) | 5 (5.55%) |
|  | Tindamba | 6 (12%) | 15 (16.66%) |
|  | Italy | 5 (10%) | 11 (12.25%) |
|  | Monou | 7 (14%) | 10 (11.11%) |
|  | Kano | 5 (10%) | 10 (11.11%) |
|  | Total | 50 | 90 |
| Dengue knowledge | Yes | 2 (4%) | 3 (3.33%) |
|  | No | 48 (96%) | 87(96.67%) |
|  | Total | 50 | 90 |
| Risky environment | Yes | 47 (94%) | 90 (100%) |
|  | No | 3 (6%) | 0 |
|  | Total | 50 | 90 |

### Dengue seroprevalence data in case and control groups

Data from Table 2 summarize the overall seroprevalence of dengue in both case and control groups as well as the type of serology detected in these patients. According to this table, the seroprevalence of dengue was lower (14%, 4/7) in malaria patients than in non-malaria patients (66.66%, 60/90). Patients from the case group were mostly found to have a secondary dengue infection (57.14%) than primary dengue infection (42.86%). The secondary dengue infection was marked by the conccurent presence of a NS1/IgM/IgG or IgM/IgG serology while primary infection was confirmed by the solely detection of IgM antibodies. Regarding the controls, primary dengue (61.66%) was more frequent than secondary dengue infection (38.34%). Patients with primary infection had a NS1 or NS1/IgM serology while those with secondary dengue infection had mostly IgM/IgG serology.

**Table 2.** Typology of dengue seropositivity in case and control groups

|  | **Cases**<br>**N=50** | **Controls**<br>**N=90** |
| --- | --- | --- |
| Dengue seropositivity rate | 7/50 (14%) | 60/90 (66.66%) |
| Primary dengue infection |  |  |
| NS1 | - | 28 |
| IgM | 3 | - |
| NS1/IgM | - | 9 |
| **Total** | **3/7 (42.86%)** | **37/60 (61.66%)** |
| Secondary dengue infection |  |  |
| IgM/IgG | 2 | 15 |
| NS1/IgM/IgG | 2 | 8 |
| **Total** | **4/7 (57.14%)** | **23/60 (38.34%)** |

### Effect of the socio-demographic parameters on DENV seropositivity

In order to identify the risk factors that might increase susceptibility to dengue infection in both malaria and non-malaria patients, the statistical association between age, gender, the dwelling place, the knowledge of dengue and the living in a risky environment with dengue seropositivity has been tested using the chi-square test. The results are presented in Tables 3 and 4 (Bold values in the P-value column are significant values at P<0.05 using the Chi-square test). As shown by these tables, no statistical association was found between dengue seropositivity and the gender, dengue knowledge, or the fact of living in a risky environment both in malaria and non-malaria patients. However, for both groups, age and the dwelling place were significantly correlated to dengue seroprevalence. As far as case group is concerned, young subjects (0-15 years old) were found to be highly susceptible to dengue compared to those of 16-30 years old (OR: 16.24; P=0.042), while some neighborhoods like Enia, Tigaza, Ngaikaida, Yademe and Italy were more risky for dengue infection (P<0.05) (Table 3). Regarding the control group, children <16 years old were also found to be highly exposed (OR: 21, P=0.0001) to dengue compared to older ones. However, only Enia, Monou and Kano were identified as risky quarters for dengue infection (Table 4).

**Table 3.** Statistical association between dengue seropositivity and socio-demographical factors in case group

| Variables |  | Dengue +<br>Malaria + | Dengue –<br>Malaria + | Odds<br>ratio<br>(OR) | OR (CI 95%) | P-value |
| --- | --- | --- | --- | --- | --- | --- |
| Gender | Male | 2(28.6%) | 18(41.8%) | 0.55 | 0.10-3.22 | 0.68 |
|  | Female | 5(71.4%) | 25(58.2%) |  |  |  |
|  | Total | 7 (100%) | 43 (100%) |  |  |  |
| Age range (years) | [0-15] | 5(71.4%) | 10(23.2%) | 16.24 | 0.80-326.1 | **0.042** |
|  | [16-30] | / | 15(34.9%) | 1 |  |  |
|  | [31-50] | 2(28.6%) | 18(41.9%) | 4.18 | 0.18-94 | 0.495 |
|  | Total | 7(100%) | 43(100%) |  |  |  |
| Dwelling Place | Enia | / | 5(11.6%) | 0 | 0.00-0.60 | **0.02** |
|  | Tigaza | / | 5(11.6%) | 0 | 0.00-0.60 | **0.02** |
|  | Ngaikaida | 1(14.3%) | 9(20.9%) | 0.04 | 0.003-0.587 | **0.03** |
|  | Yademe | / | 5(11.6%) | 0 | 0.00-0.60 | **0.02** |
|  | Mokolo | / | 2(4.7%) | 0.00 | 0.00-1.53 | 0.16 |
|  | Tindamba | 1(14.3%) | 5(11.6%) | 0.08 | 0.005-1.316 | 0.10 |
|  | Italy | / | 5(11.6%) | 0 | 0.00-0.60 | **0.02** |
|  | Monou | 5(71.4%) | 2(4.7%) | 1 |  |  |
|  | Kano | / | 5(11.6%) | 0 | 0.00-0.60 | **0.02** |
|  | Total | 7(100%) | 43(100%) |  |  |  |
| Dengue knowledge | Yes | 00 | 2(4.7%) |  |  |  |
|  | No | 7(100%) | 41(95.3%) | 0.0 | 0.0-13.77 | 0.99 |
|  | Total | 7(100%) | 43(100%) |  |  |  |
| Risky environment | Yes | 7(100%) | 40(93%) | 0.0 | 0.00-7.42 | 0.99 |
|  | No | 00 | 3(7%) |  |  |  |
|  | Total | 7(100%) | 43(100) |  |  |  |

**Table 4.** Statistical association between dengue seropositivity and socio-demographical factors in control group

| Variables |  | Dengue +<br>Malaria - | Dengue –<br>Malaria - | Odds<br>ratio<br>(OR) | OR (CI 95%) | P-value |
| --- | --- | --- | --- | --- | --- | --- |
| Gender | Male | 25(41.7%) | 11(36.7%) | 1.23 | 0.5-3.04 | 0.81 |
|  | Female | 35(58.3%) | 19(63.3%) |  |  |  |
|  | Total | 60(100) | 30(100%) |  |  |  |
| Age range (years) | [0-15] | 28(46,7%) | 2(6,7%) | 21 | 4.19-105.1 | **0.0001** |
|  | [16-30] | 12(20%) | 18(60%) | 1 |  |  |
|  | [31-50] | 20(33,3%) | 10(33,3%) | 3 | 1.046-8.605 | 0.069 |
|  | Total | 60(100%) | 30(100%) |  |  |  |
| Dwelling Place | Enia | 7(11.7%) | / | /18.33 | 0.80-416 | **0.033** |
|  | Tigaza | 3(5%) | 7(23.3%) | 0.53 | 0.081-3.5 | 0.65 |
|  | Ngaikaida | 10(16.7%) | 3(10%) | 4.167 | 0.66-26.30 | 0.187 |
|  | Yademe | 4(6.7%) | 5(16.7%) | 1 |  |  |
|  | Mokolo | / | 5(16.7%) | 0.111 | 0.0047-2.59 | 0.22 |
|  | Tindamba | 10(16.7%) | 5(16.7%) | 0.83 | 0.178- | 1 |
|  | Italy | 6(10%) | 5(16.7%) | 1.5 | 0.25-8.82 | 1 |
|  | Monou | 10(16.7%) | / | 25.67 | 1.15-59 | **0.018** |
|  | Kano | 10(16.7%) | / | 25.67 | 1.15-59 | **0.018** |
|  | Total | 60(100%) | 30(100%) |  |  |  |
| Dengue knowledge | Yes | 2(3.3%) | 1(0.3%) | 1 | 0.08-11.50 | 1 |
|  | No | 58(96.7%) | 29(96.7) |  |  |  |
|  | Total | 60(100%) | 30(100%) |  |  |  |
| Risky environment | Yes | 60(100%) | 30( 100%) |  |  |  |
|  | No | 00 | 00 |  |  |  |
|  | Total | 60(100%) | 30(100%) |  |  |  |

### Compared analysis of the hematological parameters of case and control groups

In order to gauge the influence of dengue-malaria coinfection on clinical status of patients, a compared analysis of the hematological parameters has been performed between the case and control groups. In general, co-infected subjects were found to have higher than normal levels of white blood cells (15.22 * 10^9^ cells /L >4-10 * 10^9^ cells/L), lymphocytes (9.66 * 10^9^ cells /L >0.6-4.1 * 10^9^ cells/L), and neutrophils, (13.44 * 10^9^ /L > 2-7.8 * 10^9^ / L respectively). Likewise, these subjects displayed lower than normal values of red blood cells (2.05 10^9^ / L <3.5 * 10^9^ / L), platelets (78.89* 10^9^ / L <100 * 10^9^ / L), hematocrit (29% <36%) and red blood cell volume (74 g/dL <80 g/dL). For dengue mono-infected patients, higher than normal values were recorded for the number of white blood cells (11.89 *10^9^ cells/ L), lymphocytes (8.55*10^9^ / L) and neutrophils (13.89*10^9^ / L) as well as a slight decrease of the hematocrit rate (35.56%). However, red blood cells numbers, number of platelets and the mean blood volume were relatively normal. A compared analysis of these hematological parameters revealed a significant decrease of the platelets number (P<0.001) in coinfected groups (78.89 ± 3.51 * 10^9^ / L) relative to the control groups (148.22 ± 13.94 *10^9^/L) as shown by Fig 2. Nevertheless, a non-significant decrease in red blood cells numbers, hematocrit rate the mean blood volume was found in this group.

**Fig 2 :**
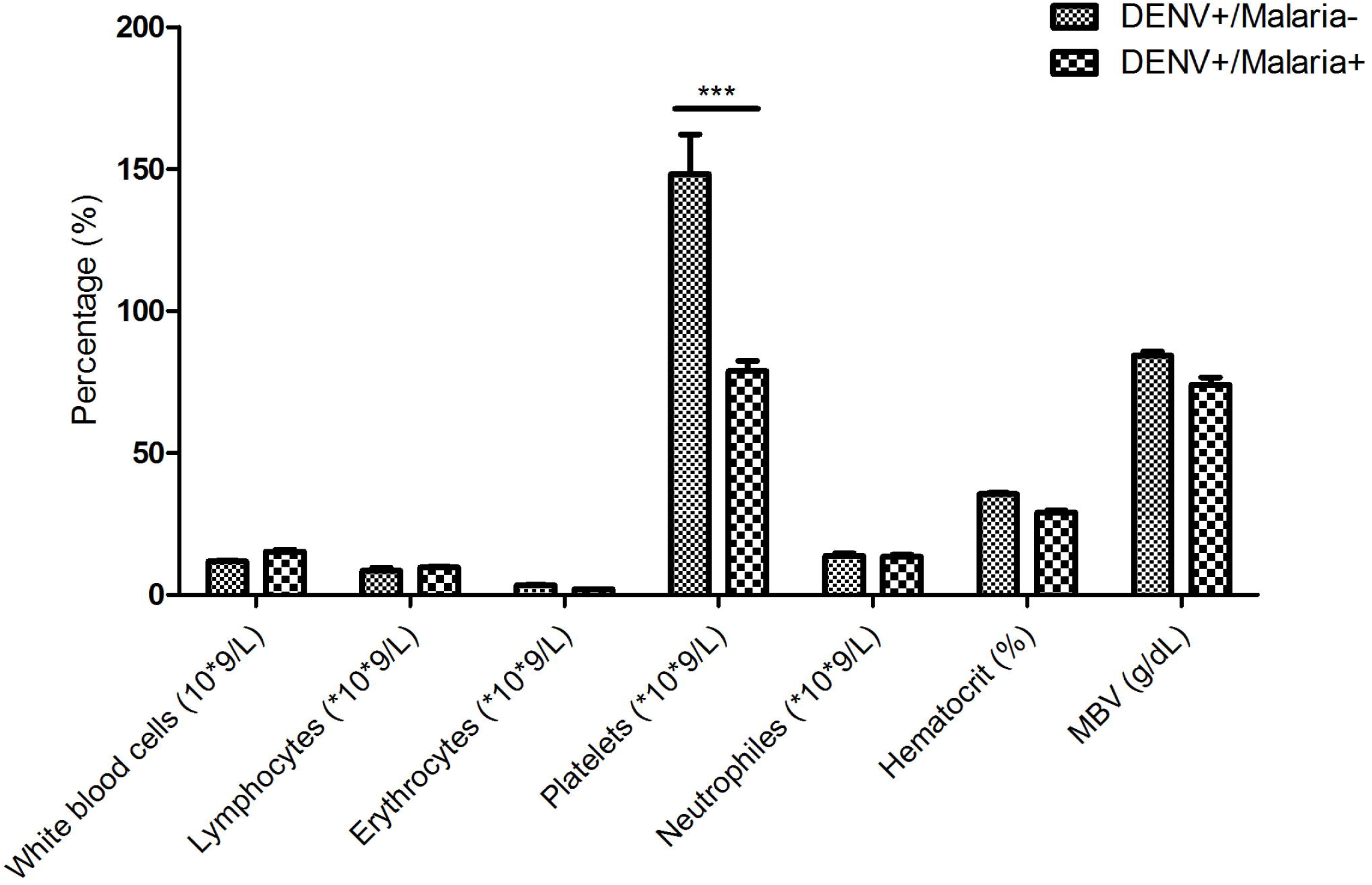
Hematological profile of dengue-malaria coinfected patients (cases) in comparison with dengue monoinfected patients (controls). ***P<0.001 when compared with the dengue- monoinfected patients

## Discussion

In Cameroon, previous studies showed a silent circulation of dengue in urban areas [26]. Moreover, concurrent infections with malaria were sporadically documented [11,18]. However, for the East region which is recognized as an endemic zone for malaria with evidence of Aedes invasion [19,20], no studies on dengue prevalence have been carried out yet. The objective of this study was to contribute to the epidemiological surveillance of this virus which remains understudied in many parts of Cameroon by a compared analysis of the seroprevalence between a dengue mono-infected cohort and dengue-malaria coinfected group consulting at the HRB. The study was carried out on patient samples collected directly at the HRB.

The serological analysis carried out on surveyed subjects revealed a dengue seroprevalence of 14% for malaria subjects against 66.6% in non-malaria patients. This result suggests that the prevalence of dengue is lower in malaria patients than in asymptomatic patients. These findings are consistent with those of Abdul-Ghani et al. who reported a coinfection rate of 4.8% versus a dengue mono-infection seroprevalence of 35.2% in outpatients from Hodeidah city in Yemen [27]. Likewise, Rao et al. found that of 745 dengue-positive cases identified in the Angul of Odisha district in India, only 3% were shown to possess dengue-malaria coinfection [28]. Also, a meta-analysis of 13 studies confirmed reduced odds of co-infected participants as compared to those with dengue mono-infection [29]. However, the coinfection rate found in this study (14%) is close of that of Yousseu et al (13%) reported in Douala (Cameroon) [14], lower than that of Mustapha et al. (44.2%) reported in Abuja (Nigeria) [30] in HIV-patients and higher than that recently documented in febrile patients from the Adamawa region (8.95%) in Cameroon by our research group [18]. Similarly, the dengue mono-infection rate found here is higher than that described by Tchuandom et al (24.8%) in blood donors recruited at the Jamot Hospital in Yaounde [17]. These discrepancies might be explained by the fact that both infections are highly endemic in the East region. Indeed, the eastern region, due to its climatic conditions [24], would be an area more favorable to the circulation of vector insects and therefore of the DENV. Particular attention to climatic factors should be taken into account in future studies to gauge its impact on the prevalence of the virus in this region. In fact, humid months with high rainfall and a temperature above 30°c have been shown to be conducive factors for high malaria-dengue coinfection rate [28]. Also, the high circulation of dengue could be due to the movement of migrants from neighboring endemic areas or the breeding sites proliferation for Aedes [30].

Considering the limitations associated with the use of rapid diagnostic tests, the underestimation of dengue cases in the present study cannot be ruled out. Other more suitable diagnostic tools with better sensitivity would certainly have made it possible to better refine the diagnosis and even to characterize the viral load of seropositive patients.

Regarding the serological analysis of dengue markers, out of the 7 cases of co-infections identified, 42.86% were mainly cases of primary infection due to the unique presence of IgM antibodies while 57.14% % were cases of secondary infection due to concomitant detection of IgG antibodies. This high proportion of IgG could indicate that the virus has been circulating for a long time in Bertoua city. These data are in line with previous studies who reported a high prevalence of malaria and dengue virus IgG in Nigeria[30]. In addition, the proportion of IgM - positive patients could be justified either by the fact that these patients were tested at a moment when the dengue infection was active and IgM antibodies were sufficient enough to be detected or they had been immunized after a recent infection. In the non-malarial population, we recorded 37 cases of patients with primary infection out of the 60 screened (61.66%) with an essentially NS1 type serology, or NS1 and IgM, which in fact reflects an active infection in these patients. The remainder (38.34%) were cases of secondary infection. This result is similar to that reported by Tchuandom et al. (2020) who found a seroprevalence of 4.5%, 12.3% and 6.1% for the NS1 antigen, IgM, and IgG antibodies respectively [17]. This also suggests that the serological profile is different between the two populations. Considering that most of the patients with secondary infection are found in the case group, it can be hypothesized that these patients were first exposed to dengue before getting malaria.

In this study, gender was not statistically associated with dengue infection, although females were found more exposed than male in the two populations. This might be due they were more represented in the study population. These results are consistent with those of a previous cameroonian study that reported an acute dengue seroprevalence of 7.7% in females versus 4.7% in males in children attending health public centers [31]. Likewise, an indian study demonstrated that of 40% of dengue-positive cases identified, women were significantly more exposed than men[32]. However, further studies have shown the contrary. Kumar et al, for example, in a gender-based study in India, reported than men were prominently more affected by dengue infection than women. The socio-cultural background which leads male to more exposed to outdoor activities with less protection for their bodies could have played a role in this exposure [33]. However, as stated by other authors the exact relationship between sociocultural background and gender-specific dengue infection rate should be deeply assessed [34]. Overall, the role of gender as risk factor for dengue infection in malaria and non-malaria patients remains controversial. Further investigations are needed to examine these variations.

Among the other demographical factors analyzed, age was statistically assciated with dengue infection in both case and control groups with subjects under 16 years of age significantly higher exposed to dengue than older participants (16-30 years) in both populations. These data are in line with previous studies carried out in Cameroon that showed a high susceptibility of infants and children to dengue infection[9,11,14,16,31]. In fact, the quality of environment in which these children live and the applying of protecting measures against mosquitoes, play an important role in the child trend of dengue infection. According to Tchuandom et al. (2019), children who live in houses where water containers are not consistently covered, insecticides not regularly sprayed or those who don’t regularly wear long-sleeved shirts have higher prevalence of dengue infection than those applying these measures [31].

Regarding places of residence, the Enia Tigaza, Ngaikada, Yademe, Italy and Kano neighborhoods were more associated with dengue infection in coinfected patients (P<0.05) while the Enia, Monou and Kano districts were the only significant sources of contamination for dengue-monoinfected patients. Given that dengue prevalence was higher in monoinfected patients, it could be suggested that Enia, Monou and Kano districts which belong to Bertoua II sub-division are the main disseminating areas for DENV and thus for Aedes propagation. In a geographical study on distribution and prevalence of Aedes species through Cameroon, Tedjou et al reported a seroprevalence of 99.1% for *Ae. albopictus* vs 0.9% for *Ae. aegypti* in Bertoua downtown while the vectors were present at 97.97% vs 2.03% rates respectively in Bertoua suburban areas [20].

With regard to the haematological parameters, the presence of dengue in the 2 populations showed a variation in the haematological parameters. The number of lymphocytes, neutrophils and white blood cells increased in both mono-infected and co-infected compared to normal values, but this increase was not significant between the two groups. The number of red blood cells and the hematocrit level were considerably lower in the co-infected than in the mono-infected with an insignificant variation between the two groups. Likewise, the mean corpuscular volume was slightly lower in co-infected than in mono-infected. However, a significant decrease in the number of platelets was observed in co-infected patients compared to mono-infected patients (78.89 ± 3.51 * 10^9^ / L vs 148.22 ± 13.94 *10^9^/L). There may be a few reasons for the thrombocytopenia observed in co-infected such as platelet consumption by activated macrophages during phagocytosis, increased peripheral sequestration, or activation of the complement system as previously stated [35]. On contrary to these data, a meta-analysis study by Kotepui et al. (2019) demonstrated a higher odds of platelets among coinfected patients relative to dengue-monoinfected patients [29]. Moreover, some studies did not find any significant difference in platelets levels between both groups [36]. However, Barua and Gill in 2016 reported a significantly lower platelet value in co-infected patients compared to those uni- infected with dengue (47,587 versus 76,422 / mm3) [37].

One of the limitations of this study could be the sample size which, in the future, must be increased to obtain more representative data. Due to resource constraints, it has not been possible to confirm positive IgM by plaque reduction neutralization tests, and serotypes of NS1 positive cases. The need to extend sampling throughout the year instead of just 2 months will allow us to better identify the periods at risk of dengue infection in the Easr region and to analyze the impact of the climate.

## Conclusion

At the end of this work aiming to analyze the circulation of the dengue virus in malaria patients at the BRH and to compare it with a non-malaria cohort, we can retain that malaria patients have statistically fewer cases of dengue than non-malaria patients. The majority of these cases are dengue secondary infection, whereas in the mono-infected group it is much more of a primary infection. In addition, in the two populations, gender was not statistically associated with seropositivity to dengue, but age and the fact of living in certain neighborhoods of the city of Bertoua would statistically increase the chances of being exposed to dengue. Regarding haematological parameters, we noted that the decrease in blood platelets is the flagship hematological modification that can lead a malaria patient to be suspected of a case of dengue, even though alterations in white blood cells, red blood cells and hematocrit can be noted. The results of this study point the endemic character of dengue infection in East-Cameroon. Further investigations are necessary to characterize the circulating serotypes and even follow up patients with mono- or co-infection for complications.

## Data Availability

Data are available upon request to the corresponding author

## Acknowledgments

We express our sincere thanks to all the volunteers who participated to this study. Also, the technical assistance of the personal from the Laboratory of the BRH is greatly acknowledged. The study was partly possible thanks to an research grant (F/6125-1) from International Foundation for Science, which helped us to acquire Dengue kits. The funders had no role in study design, data collection and analysis, decision to publish, or preparation of the manuscript

## Declaration of Interests

The authors of this manuscript have no competing interests to disclose.

## Notes

### Competing Interest Statement

The authors have declared no competing interest.

### Funding Statement

The study was partly possible thanks to a research grant (F/6125-1) from International Foundation for Science, which helped us to acquire Dengue kits.

### Author Declarations

This study protocol was approved by the National Ethics Committee for Research on Human Health Ethical Clearance Number, 2020/01/58/ CE/CNERSH/SP. Permission for sample collection at BRH was granted by the Regional Delegate for Public Health of the East region (Authorization Number 622/L/MINSANTE/SG/DRSPE/BFP).

